# Comparing children’s night cough with wheeze: phenotypic characteristics, healthcare use and treatment

**DOI:** 10.1101/2022.07.05.22277192

**Authors:** Maria Christina Mallet, Rebeca Mozun, Cristina Ardura-Garcia, Eva SL Pedersen, Maja Jurca, Philipp Latzin, LUIS study group, Alexander Moeller, Claudia E. Kuehni

## Abstract

Population-based studies of children presenting with dry night cough alone compared with those who also wheeze are few and inconclusive.

Luftibus in the school is a population-based study of schoolchildren conducted between 2013–2016 in Zurich, Switzerland. We divided children into four mutually exclusive groups based on reported dry night cough (‘cough’) and wheeze and compared parent-reported symptoms, comorbidities and exposures using multinomial regression, FeNO using quantile regression, spirometry using linear regression and healthcare use and treatments using descriptive statistics.

Among 3457 schoolchildren aged 6–17 years, 294 (9%) reported ‘cough’, 181 (5%) reported ‘wheeze’, 100 (3%) reported ‘wheeze and cough’ and 2882 (83%) were ‘asymptomatic.’ Adjusting for confounders in a multinomial regression, children with ‘cough’ reported more frequent colds, rhinitis and snoring than ‘asymptomatic’ children; children with ‘wheeze’ or ‘wheeze and cough’ more often reported hay fever, eczema and parental histories of asthma. FeNO and spirometry were similar among ‘asymptomatic’ and children with ‘cough,’ while children with ‘wheeze’ or ‘wheeze and cough’ had higher FeNO and evidence of bronchial obstruction. Children with ‘cough’ used healthcare less often than those with ‘wheeze,’ and they attended mainly primary care. Twenty-two children (7% of those with ‘cough’) reported a physician diagnosis of asthma and used inhalers. These had similar characteristics as children with wheeze.

Our representative population-based study suggests only a small subgroup (7%) of schoolchildren reporting dry night cough without wheeze have features typical of asthma, yet the majority (93%) should be investigated for alternative aetiologies, particularly upper airway disease.

**Take home message:** Our population-based study found children with night cough alone clearly differ from those with wheeze, suggesting different aetiologies and pathophysiology. Yet, a small subgroup (7%) has features of asthma and may benefit from specific work-up.

## Introduction

Cough is a common symptom affecting children’s and their families’ quality of life and burdening healthcare systems [1]. Respiratory tract infections are the most common cause of cough, followed by asthma which is characterised by cough and wheeze [2–4]. Cough is also present in rare lung diseases, such as cystic fibrosis, primary ciliary dyskinesia or interstitial lung diseases [5]. In absence of an obvious respiratory infection or an underlying severe disease, many children have so-called non-specific cough [2], which is a diagnostic conundrum for physicians and a source of parental worries [6]. Underlying causes of non-specific cough include environmental exposures, (*e.g*. tobacco smoke, allergens), ear, nose and throat (ENT) problems, post-infectious cough or atypical asthma [5, 7–10]. Although wheeze is a key symptom of asthma [11], some researchers proposed that children can have ‘cough variant asthma’ without audible wheeze [12], yet others fear this construct may lead to asthma overdiagnosis, unnecessary treatments, side effects and increased costs [6, 13–16].

Previous studies on non-specific cough were small, included selected participants from specialist clinics [17–19] or relied only on self-reported data [14, 20, 21]. Few were population-based and included information on measurable asthma traits [22–25]. Only two studies distinguished between children with wheeze alone or cough alone from those with both symptoms [22, 26]. Our Luftibus in the school (LUIS) study of unselected schoolchildren obtained information on parent-reported wheeze, cough, upper respiratory symptoms, environmental exposures, and healthcare visits; we also measured fractional exhaled nitric oxide (FeNO) and lung function, which are important asthma-related traits [27]. We determined how frequently schoolchildren reported dry night cough (henceforth ‘cough’) and wheeze alone and in combination and how these groups differed. Our underlying motivation was gaining insight into the aetiology of non-specific cough and investigating possibilities that some children may have a variant form of asthma. We compared four groups (‘cough’, ‘wheeze’, ‘wheeze and cough’ and ‘asymptomatic’) of children with respect to sociodemographic and environmental information, family history, parent-reported symptoms and comorbidities, FeNO and lung function, healthcare utilisation and asthma diagnosis and treatment.

## Methods

### Study design and population

We conducted LUIS from 2013–2016 in the canton of Zurich, Switzerland, as a cross-sectional population-based study of 6–17-year-old schoolchildren (ClinicalTrials.gov: NCT03659838) [28]. All schools were invited and whole classes recruited. Parents completed questionnaires about respiratory symptoms, diagnoses and treatments, lifestyle and household characteristics. Trained lung function technicians measured FeNO and performed spirometry in a mobile bus with lung function equipment. The ethics committee of the canton of Zurich approved the study (KEK-ZH-Nr: 2014-0491); written informed consent was obtained from parents and verbal and, where appropriate, written consent was obtained from children.

### Outcomes: definition of cough and wheeze

Previous studies used different definitions to assess non-specific cough, such as persistent cough, recurrent cough, dry night cough or cough apart from cold [14, 20–24, 26, 29]. For our study, we used the question from the International Study of Asthma and Allergy in Childhood (ISAAC): “In the last 12 months has your child had a dry cough at night, apart from a cough associated with a cold or a chest infection?” (Table S1). We also used a question from ISAAC to assess wheeze: “Did your child have wheezing or whistling in the chest in the past 12 months?” [30] Based on answers to these two questions, we defined four mutually exclusive groups: ‘cough,’ ‘wheeze,’ ‘wheeze and cough’ and ‘asymptomatic.’

### Characteristics: socioeconomic, environmental, comorbidities, healthcare utilisation and treatment

Based on a literature search, we selected a range of exposures associated with cough, wheeze or asthma, including a) sociodemographic factors [sex, age, country of origin, socioeconomic status (Swiss socioeconomic position index, SSEP)]; b) environmental exposures (urbanisation degree, household pets, siblings, parental smoking); c) comorbidities [body mass index (BMI), frequency of colds, family history of asthma and chronic cough, personal history of atopy (eczema, hay fever)] and ENT problems, such as rhinitis apart from colds, otitis media, snoring (apart from colds and almost every night) and adenotonsillectomy. We also investigated potential triggers for cough or wheeze, asking specifically about exercise, respiratory infections, aeroallergens (house dust, pollen and pets), physical factors (cold air/fog, laughter, weather/temperature changes) and certain foods and drinks [14, 21, 22, 31, 32]. We include the questions in Table S1. We also compared how different types of healthcare utilisation, frequency of physician asthma diagnosis ever and asthma treatment in the past 12 months differed across the four groups.

### Measurements: FeNO and lung function

Trained technicians measured FeNO [expressed in parts per billion (ppb)] with the single-breath online method according to the American Thoracic Society (ATS)/European Respiratory Society (ERS) recommendations [33] using a chemiluminescence analyser (CLD88, Eco Medics, Dürnten, Switzerland). Spirometry was performed using Masterlab, Jaeger, Würzburg, Germany according to ATS/ERS guidelines and paediatric pulmonologists did a post hoc quality control of flow-volume curves [34]. Using Global Lung Initiative (GLI) reference values [35], we derived z-scores for forced vital capacity (FVC), forced expiratory volume in the first second (FEV1) and forced expiratory flow between the 25% and 75% of the FVC (FEF 25–75). We also calculated the FEV1/FVC ratio.

### Statistical analysis

We compared sociodemographic information, environmental factors, family history and symptoms between the four groups of children by calculating proportions, means with standard deviations (SD), and medians with interquartile range (IQR). We first tested differences between groups using chi-square tests for categorical variables and the Kruskal-Wallis test for non-normally distributed continuous data and in a second step, using multinomial logistic regression with ‘asymptomatic’ as the reference group. We report unadjusted and adjusted relative risk ratios (RRR) and 95% confidence intervals (95% CI). For our final model, we included all variables that had a p-value of <0.05 in univariable analysis, and then we applied stepwise backward selection. We performed likelihood ratio tests between the full model and reduced models and only kept variables if the likelihood ratio test had a p<0.05. Sex and age were kept a priori.

We then investigated how FeNO and lung function parameters differed between groups using regression analyses. For FeNO, we used quantile regression analysis because FeNO was not normally distributed. We adjusted for age, sex, hay fever, inhaled corticosteroids (ICS), use of beta-2 receptor antagonist and smoking—factors potentially influencing FeNO [36–38]. We also computed four linear regression models with z-scores of FEV1, FVC, FEV1/FVC, and FEF25-75 as outcome and the four groups (‘cough,’ ‘wheeze,’ ‘wheeze and cough’ and ‘asymptomatic’) as explanatory variable, and we adjusted for BMI z-score. We compared types of healthcare utilisation, frequency of physician asthma diagnosis ever and asthma treatment in the past 12 months by calculating proportions and 95% CI.

In a post hoc analysis to investigate accurate asthma diagnosis and treatment, we compared characteristics of 22 children with ‘cough’ who reported physician diagnosis of asthma and use of asthma inhalers [short acting bronchodilators (SABA) or ICS] in the past 12 months with children reporting ‘wheeze,’ ‘wheeze and cough’ and the remaining 272 children with ‘cough.’

We recoded missing values for symptom questions as ‘no’; we assumed absent or mild symptoms when parents had not answered with ‘yes’ (Table S1). We used STATA (Version 15.1, StataCorp) for statistical analysis.

## Results

We included 3457 children (50% male) from 37 schools (Figure S1) with a median age of 13 years (range 6–17) (Table 1). Parents reported children with dry night cough alone (‘cough’) for 294 (9%); wheeze alone (‘wheeze’) for 181 (5%); ‘wheeze and cough’ for 100 (3%); and neither symptom (‘asymptomatic’) for 2882 (83%) (Figure 1A). Twenty-two children with ‘cough’ (7% of 294) were diagnosed with asthma by physicians and reported using asthma inhalers (SABA or ICS) in the past 12 months (Figure 1B).

**Figure 1.**
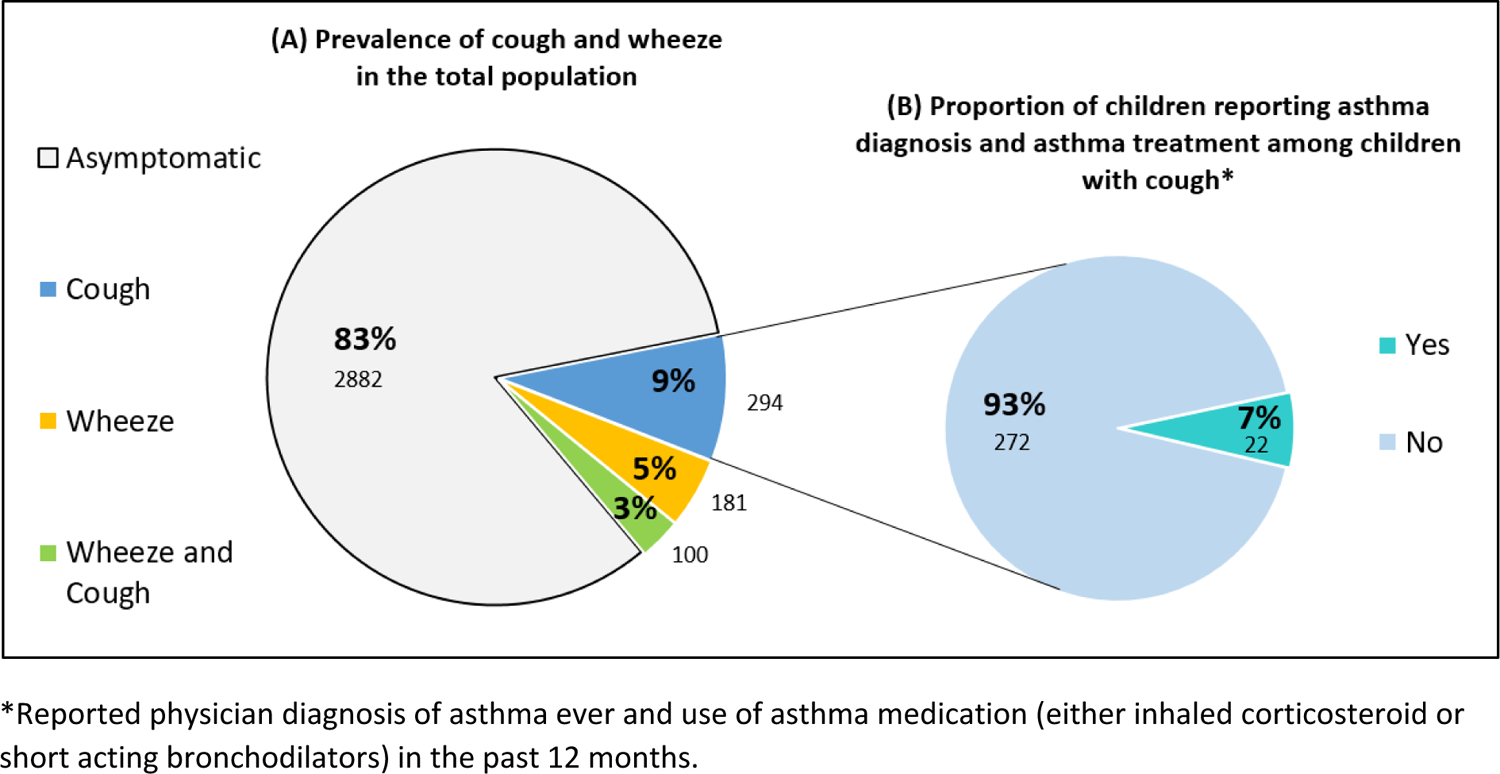
**(A)** Reported prevalence of cough, wheeze, wheeze and cough, and asymptomatic in the LuftiBus in the School study (N=3457) **(B)** Proportion of children with physician-diagnosed asthma and asthma medication among children with cough (N= 294)

**Table 1:** Characteristics of asymptomatic children, children with cough alone (‘cough’), wheeze alone (‘wheeze’), and ‘wheeze and cough’ in the LuftiBus in the School study (N=3457)

|  | Total<br>(N=3457)<br>n (%) | Asymptomatic<br>(N=2882)<br>n (%) | Cough<br>(N=294)<br>n (%) | Wheeze<br>(N=181)<br>n (%) | Wheeze<br>and cough<br>(N=100)<br>n (%) | p-value |
| --- | --- | --- | --- | --- | --- | --- |
| **Sociodemographic factors** |  |  |  |  |  |  |
| Male sex | 1719 (50) | 1446 (50) | 132 (45) | 85 (47) | 56 (56) | 0.162 |
| Median age in years (IQR) | 13 (10–14) | 13 (10–14) | 12 (9–14) | 13 (11–14) | 12 (9–14) | 0.011<sup>1</sup> |
| Child, country of birth CH | 3042 (89) | 2527 (88) | 266 (91) | 167 (93) | 82 (82) | 0.019 |
| Swiss SEP median° (IQR) | 69.7<br>(62.2–76.6) | 69.9<br>(62.5–76.9) | 68.9<br>(59.3–75.1) | 70.2<br>(62.7–76.1) | 68.8<br>(62.5–75.1) | 0.036<sup>1</sup> |
| **Environmental exposures** |  |  |  |  |  |  |
| Large urban area | 1674 (48) | 1400 (49) | 157 (53) | 77 (43) | 40 (40) | 0.040 |
| Household pets | 1469 (43) | 1244 (44) | 105 (36) | 82 (46) | 38 (38) | 0.055 |
| Maternal smoking | 571 (17) | 483 (17) | 52 (18) | 17 (9) | 19 (19) | 0.055 |
| Paternal smoking | 757 (22) | 634 (22) | 68 (24) | 36 (20) | 19 (20) | 0.746 |
| **Parental history of** |  |  |  |  |  |  |
| Asthma | 486 (14) | 347 (12) | 43 (15) | 69 (39) | 27 (27) | <0.001 |
| Chronic cough | 180 (6) | 130 (5) | 20 (7) | 20 (11) | 10 (11) | <0.001 |
| **Symptoms and comorbidities/clinical factors** |  |  |  |  |  |  |
| Median BMI z-score* (IQR) | -0.01<br>(-0.70–0.80) | -0.03<br>(-0.71–0.76) | 0.04<br>(-0.65–0.86) | 0.26<br>(-0.71–0.96) | 0.29<br>(-0.4–1.00) | 0.005<sup>1</sup> |
| Eczema<sup>§</sup> | 249 (7) | 184 (6) | 23 (7) | 30 (17) | 13 (13) | <0.001 |
| Rhinitis<sup>§</sup> | 1048(30) | 726 (25) | 139 (47) | 115 (64) | 68 (68) | <0.001 |
| Hay fever | 766 (22) | 549 (19) | 74 (25) | 94 (52) | 49 (49) | <0.001 |
| Frequent colds (>3/year)<sup>§</sup> | 587 (17) | 407 (14) | 91 (31) | 50 (28) | 39 (39) | <0.001 |
| Otitis media at least once<sup>§</sup> | 333 (10) | 262 (9) | 36 (12) | 13 (7) | 22 (22) | <0.001 |
| Snoring<sup>§#</sup> | 888 (26) | 697 (24) | 105 (36) | 53 (29) | 33 (33) | <0.001 |
| **Triggers for wheeze or cough<sup>§</sup>** |  |  |  |  |  |  |
| Exercise | 614 (18) | 373 (13) | 85 (29) | 94 (52) | 62 (62) | <0.001 |
| Dust | 217 (6) | 106 (4) | 38 (13) | 40 (22) | 33 (33) | <0.001 |
| Pollen | 383 (11) | 208 (7) | 51 (17) | 78 (43) | 46 (46) | <0.001 |
| Pets | 128 (4) | 54 (2) | 15 (5) | 37 (20) | 22 (22) | <0.001 |
| Cold air | 334 (10) | 200 (7) | 48 (16) | 47 (26) | 39 (39) | <0.001 |
| Food and drink | 100 (3) | 64 (2) | 17 (6) | 12 (7) | 7 (7) | <0.001 |
| Laughter | 284 (8) | 193 (7) | 37 (13) | 34 (19) | 20 (20) | <0.001 |
| Weather/temperature change | 267 (8) | 159 (6) | 52 (18) | 27 (15) | 29 (29) | <0.001 |
| Colds/infections | 2138 (62) | 1703 (59) | 214 (73) | 136 (75) | 85 (85) | <0.001 |

IQR: Interquartile range; CH: Switzerland; SEP: Socioeconomic position; ° Swiss SEP ranges from 0-lowest (worse) to 100-highest (better); \*BMI: Body Mass Index, we calculated BMI z-scores using references values from the World Health Organisation; § refer to symptoms in the past 12 months ; # snoring includes those who snore sometimes without cold or almost every night; p-values calculated using chi square test; <sup>1</sup> p-values calculated using Kruskal-Wallis

### Characteristics of children with ‘cough,’ ‘wheeze,’ ‘wheeze and cough’ and ‘asymptomatic’

The four groups differed in regard to environmental exposures, parental history, symptoms and symptom triggers (Tables 1 & S2, Figure 2).

**Figure 2:**
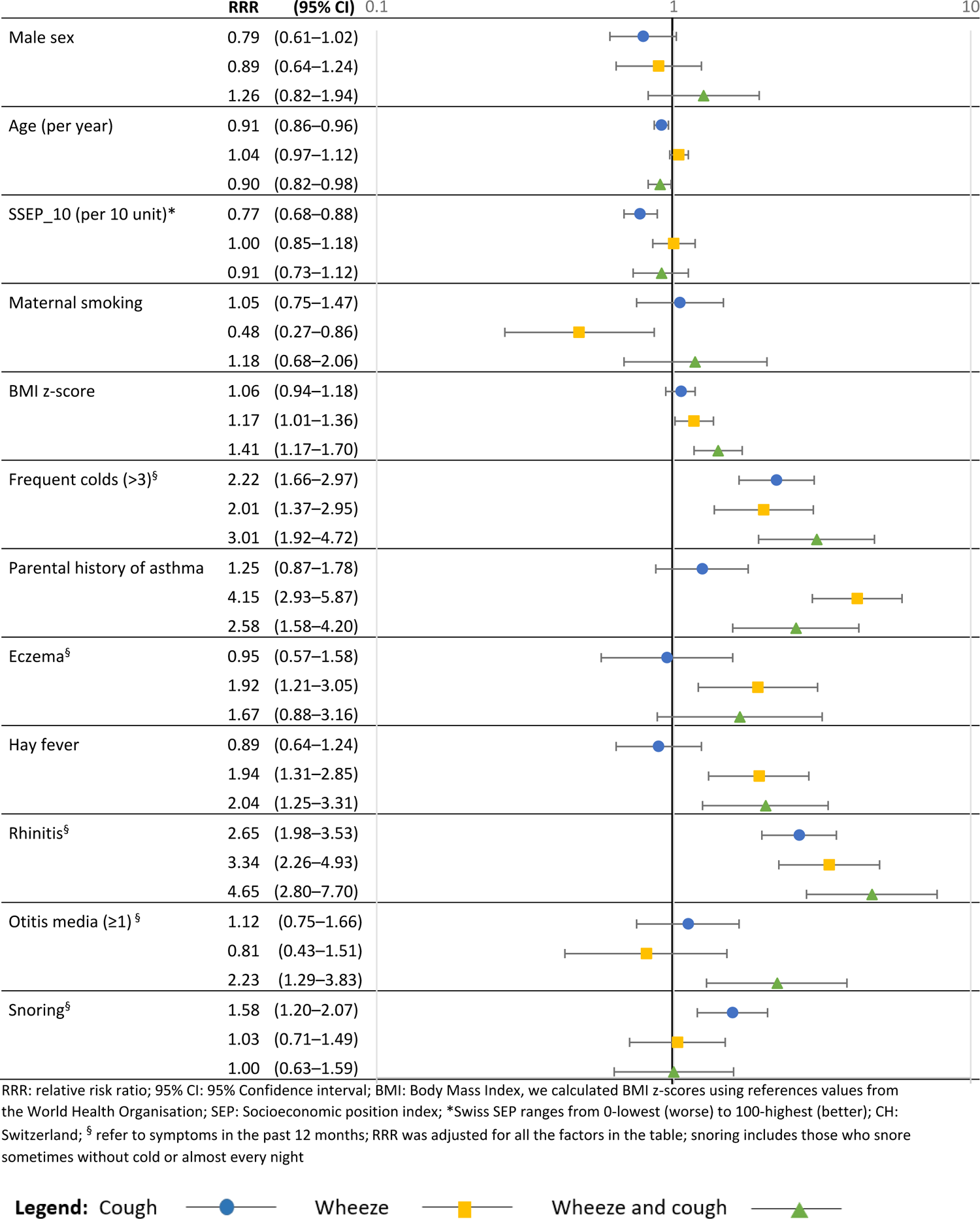
Adjusted relative risk ratios of factors associated with cough (N=294), wheeze (N=181) and wheeze and cough (N=100) compared to asymptomatic children (N=2882) (reference group)

Children with ‘cough’ were less often male (RRR 0.79, 95%CI 0.61–1.02) and younger (RRR 0.91, 95%CI 0.86–0.96) with lower SSEP (RRR 0.77, 95%CI 0.68–0.88) than ‘asymptomatic’ children. They also reported more colds in the past 12 months (RRR 2.22, 95%CI 1.66–2.97), more rhinitis apart from colds (RRR 2.65, 95%CI 1.98–3.53) and more snoring (RRR 1.58, 95%CI 1.20–2.07). There were no differences between children with ‘cough’ and ‘asymptomatic’ children regarding tobacco exposure and personal or family history of atopic diseases (Figure 2, Table S2).

Children either with ‘wheeze’ or ‘wheeze and cough’ reported more frequent colds and rhinitis than asymptomatic children, and there were strong associations with parental history of asthma (RRR 4.15, 95%CI 2.93–5.87 for ‘wheeze’ and RRR 2.58, 95%CI 1.58–4.20 for ‘wheeze and cough’) and a personal history of hay fever (RRR 1.94, 95%CI 1.31–2.85 for ‘wheeze’ and RRR 2.04, 95%CI 1.25–3.31 for ‘wheeze and cough’). BMI was associated with both outcomes—the association was stronger for ‘wheeze and cough.’ A report of otitis media was associated with ‘wheeze and cough’ (RRR 2.23, 95%CI 1.29–3.83).

All trigger factors for cough or wheeze were reported more often among children with ‘wheeze’ or ‘wheeze and cough’ when compared with children with ‘cough’ (Table 1). House dust as a trigger was reported more often for children with ‘wheeze and cough’ (33%) than for children with ‘wheeze’ (22%, p=0.046), and some physical triggers also differed between the two groups [cold air: 39% for ‘wheeze and cough’ vs. 26% for ‘wheeze’ (p=0.023); weather/temperature change, 29% vs. 15% (p=0.005)].

### FeNO and lung function tests

Children with ‘cough’ had similar FeNO levels (median 11.5 ppb, IQR 6.6–20.6) as asymptomatic children (median 11.8, IQR 7.0–19.9) (Table 2), which we confirmed in the adjusted regression model (Figure 3, Table S3). FEV1, FEV1/FVC and FEF 25-75 z-scores were comparable among children with ‘cough’ and ‘asymptomatic’ children (Table 2, Figure 4, Table S3). FVC was higher among children with ‘cough’ (+0.170 z-scores, 95% CI 0.020– 0.320), even after adjustment for BMI (+0.143 z-scores, 95% CI 0.000–0.286).

**Figure 3:**
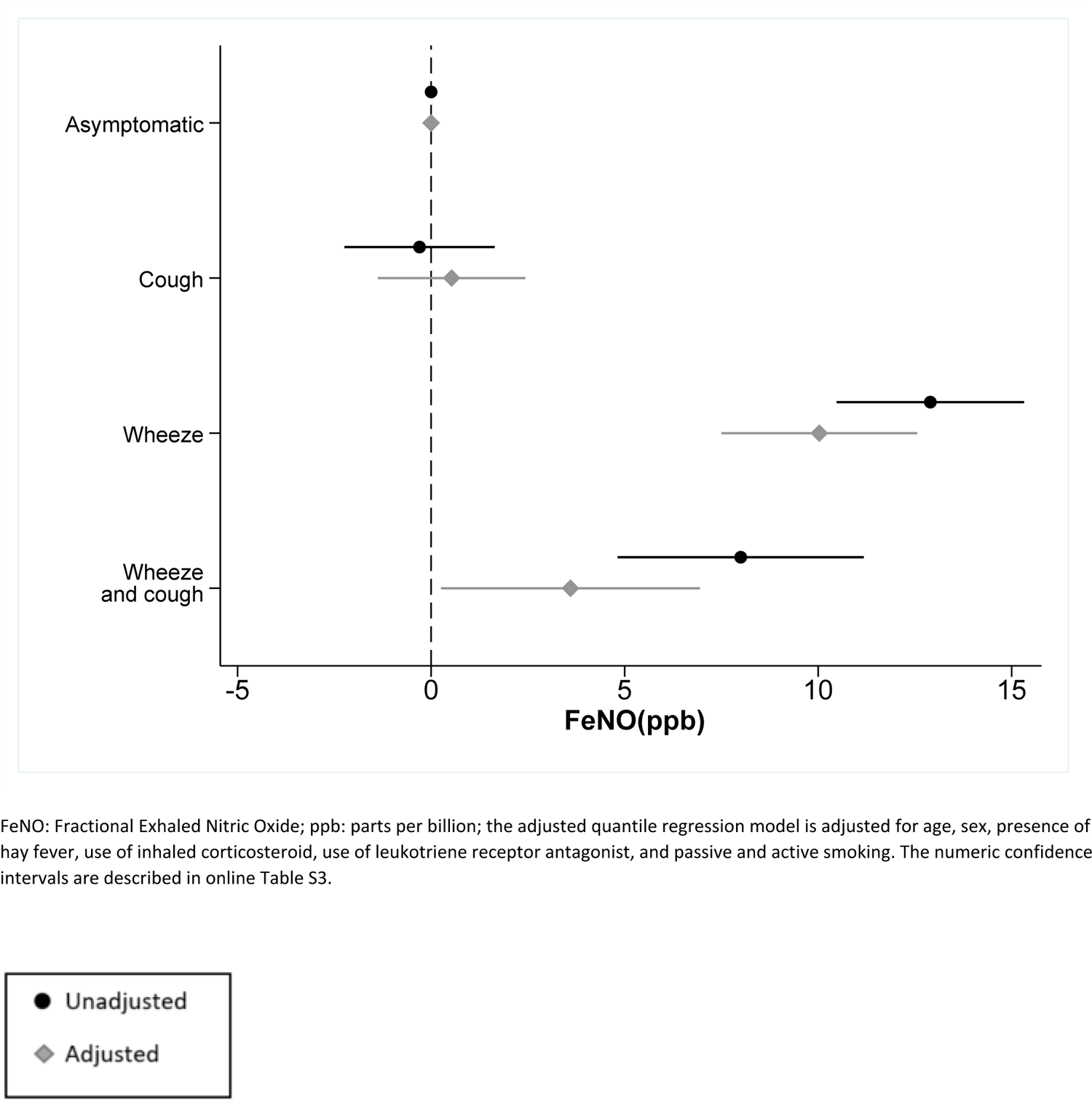
Association of fractional exhaled nitric oxide (FeNO) among children with cough (N=251), wheeze (N=156), and wheeze and cough (N=88) compared to asymptomatic children (N=2535) (reference group)

**Figure 4:**
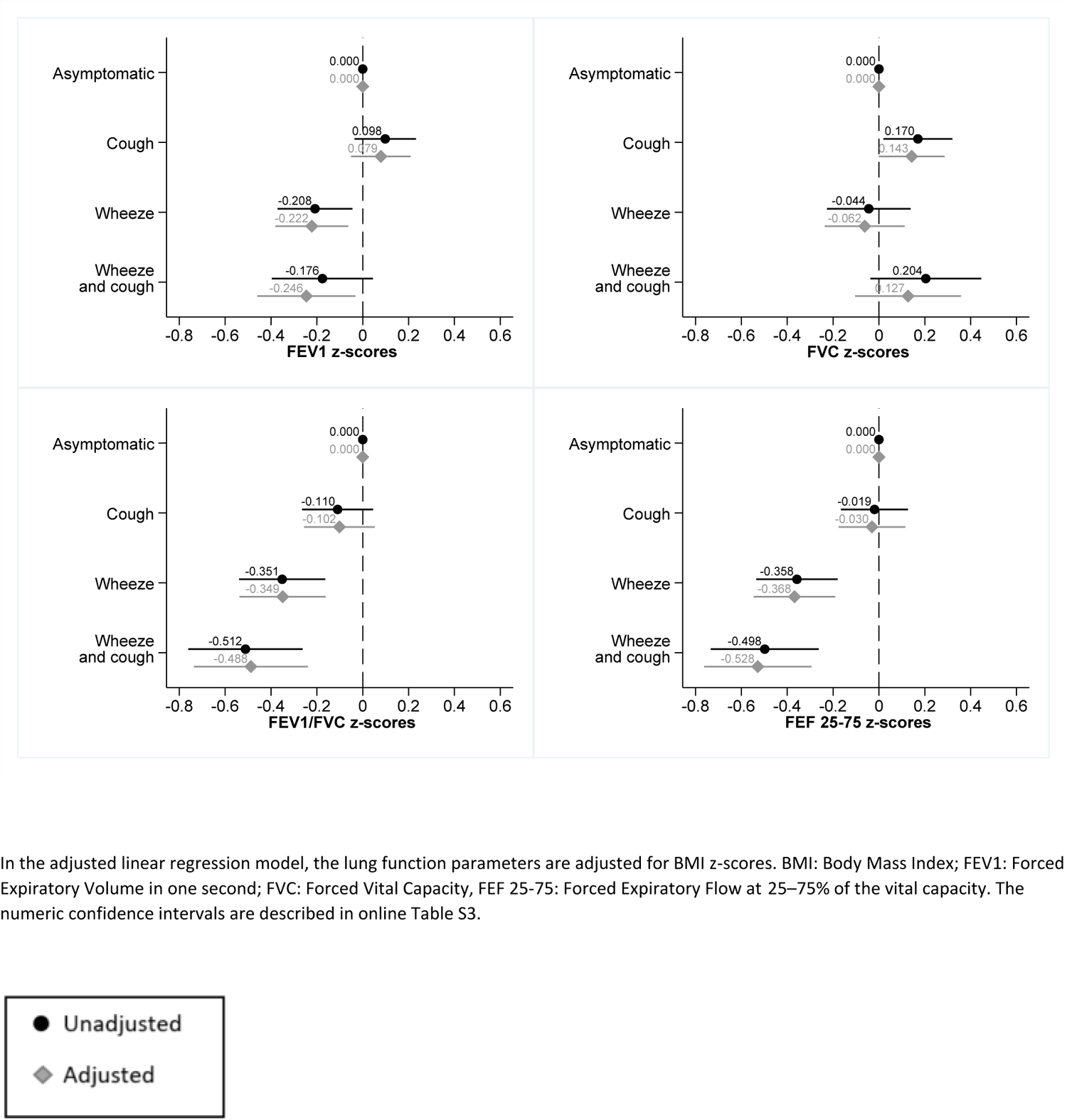
Spirometry z-scores for children with cough (N=230), wheeze (N=150), and wheeze and cough (N=80) compared with asymptomatic children (N=2343) (reference group)

**Table 2:**
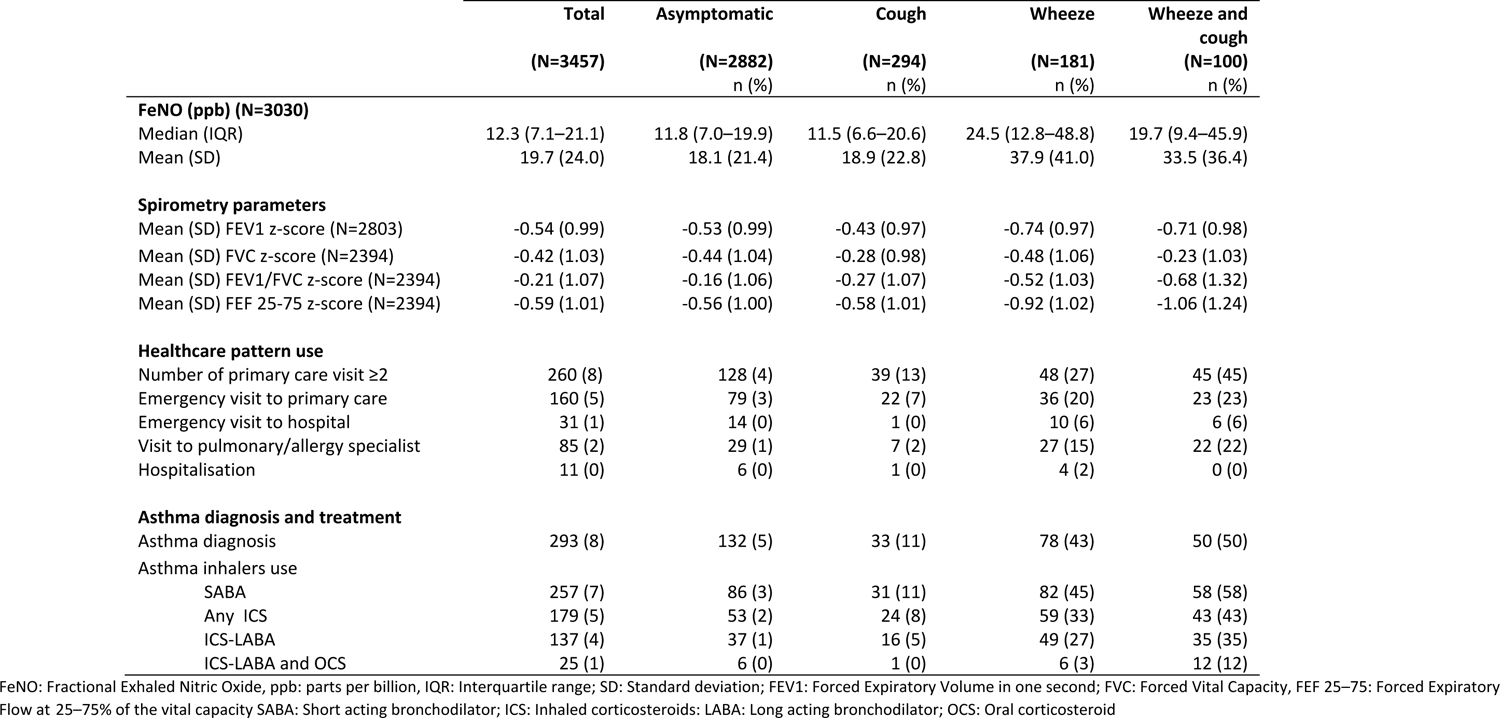
Comparison of measured traits (FeNO and spirometry), healthcare pattern, asthma diagnosis, and treatment between asymptomatic children, children with cough, wheeze, and wheeze and cough in the LuftiBus in the School study (N=3457)

|  | Total<br>(N=3457) | Asymptomatic<br>(N=2882)<br>n (%) | Cough<br>(N=294)<br>n (%) | Wheeze<br>(N=181)<br>n (%) | Wheeze and<br>cough<br>(N=100)<br>n (%) |
| --- | --- | --- | --- | --- | --- |
| **FeNO (ppb) (N=3030)** |  |  |  |  |  |
| Median (IQR) | 12.3 (7.1–21.1) | 11.8 (7.0–19.9) | 11.5 (6.6–20.6) | 24.5 (12.8–48.8) | 19.7 (9.4–45.9) |
| Mean (SD) | 19.7 (24.0) | 18.1 (21.4) | 18.9 (22.8) | 37.9 (41.0) | 33.5 (36.4) |
| **Spirometry parameters** |  |  |  |  |  |
| Mean (SD) FEV1 z-score (N=2803) | -0.54 (0.99) | -0.53 (0.99) | -0.43 (0.97) | -0.74 (0.97) | -0.71 (0.98) |
| Mean (SD) FVC z-score (N=2394) | -0.42 (1.03) | -0.44 (1.04) | -0.28 (0.98) | -0.48 (1.06) | -0.23 (1.03) |
| Mean (SD) FEV1/FVC z-score (N=2394) | -0.21 (1.07) | -0.16 (1.06) | -0.27 (1.07) | -0.52 (1.03) | -0.68 (1.32) |
| Mean (SD) FEF 25-75 z-score (N=2394) | -0.59 (1.01) | -0.56 (1.00) | -0.58 (1.01) | -0.92 (1.02) | -1.06 (1.24) |
| **Healthcare pattern use** |  |  |  |  |  |
| Number of primary care visit ≥2 | 260 (8) | 128 (4) | 39 (13) | 48 (27) | 45 (45) |
| Emergency visit to primary care | 160 (5) | 79 (3) | 22 (7) | 36 (20) | 23 (23) |
| Emergency visit to hospital | 31 (1) | 14 (0) | 1 (0) | 10 (6) | 6 (6) |
| Visit to pulmonary/allergy specialist | 85 (2) | 29 (1) | 7 (2) | 27 (15) | 22 (22) |
| Hospitalisation | 11 (0) | 6 (0) | 1 (0) | 4 (2) | 0 (0) |
| **Asthma diagnosis and treatment** |  |  |  |  |  |
| Asthma diagnosis | 293 (8) | 132 (5) | 33 (11) | 78 (43) | 50 (50) |
| Asthma inhalers use |  |  |  |  |  |
| SABA | 257 (7) | 86 (3) | 31 (11) | 82 (45) | 58 (58) |
| Any ICS | 179 (5) | 53 (2) | 24 (8) | 59 (33) | 43 (43) |
| ICS-LABA | 137 (4) | 37 (1) | 16 (5) | 49 (27) | 35 (35) |
| ICS-LABA and OCS | 25 (1) | 6 (0) | 1 (0) | 6 (3) | 12 (12) |

FeNO: Fractional Exhaled Nitric Oxide, ppb: parts per billion, IQR: Interquartile range; SD: Standard deviation; FEV1: Forced Expiratory Volume in one second; FVC: Forced Vital Capacity, FEF 25–75: Forced Expiratory Flow at 25–75% of the vital capacity SABA: Short acting bronchodilator; ICS: Inhaled corticosteroids; LABA: Long acting bronchodilator; OCS: Oral corticosteroid

Children with ‘wheeze’ and ‘wheeze and cough’ had increased FeNO [median 24.5 ppb (IQR 12.8–48.8) and 19.7 ppb (IQR 9.4–45.9), respectively] when compared with ‘asymptomatic’ and children with ‘cough.’ Differences remained after adjusting for potential confounders in the quantile regression (Figure 3, Table S3). Children with ‘wheeze’ and ‘wheeze and cough’ also had lower z-scores for FEV1, FEV1/FVC and FEF25-75 (Figure 4 and Table S3).

### Healthcare utilisation

Children with ‘wheeze and cough’ used healthcare more than children with ‘wheeze’ or ‘cough’ (Table 2, Figure S2A). Only 13% of children with ‘cough’ reported two or more visits to paediatricians and 7% required emergency consultations in primary care in the last 12 months. Visits to pulmonologists or allergologists were mainly reported by children with ‘wheeze’ (15%) or ‘wheeze and cough’ (22%). Hardly any children (<1%) with ‘cough’ reported emergency hospital visits or hospitalisations for respiratory problems.

### Asthma diagnosis and treatment

A physician diagnosis of asthma ever was reported among 8% of children overall (Table 2, Figure S2B). Half of the children with ‘wheeze and cough’ and 43% of those with ‘wheeze’ reported physician diagnosis of asthma compared with 11% of children with ‘cough’ and 5% of asymptomatic children. Asthma inhalers were prescribed most often for children with ‘wheeze and cough’ (SABA: 58%; any ICS: 43%) and ‘wheeze’ (SABA: 45%; any ICS: 33%); 11% of children with ‘cough’ received SABA and 8% ICS in the last year.

### Subgroup analysis: children with ‘cough’ who were diagnosed and treated for asthma

For a small fraction (7%, N=22) of children with ‘cough,’ parents reported a diagnosis of asthma and current treatment with SABA or ICS (Figure 1B). These children resembled in all aspects children with ‘wheeze’ or ‘wheeze and cough’ (Table S4) and were distinct from other children reporting ‘cough’ alone. They frequently had positive family histories of asthma (43%), personal histories of atopic dermatitis (14%) and hay fever (50%); they reported respiratory symptoms triggered by exercise (59%) or pollen (41%). Their median (IQR) FeNO values [24.7 (8.8–52.7) ppb] and mean (SD) lung function z-scores [FEV1: −0.78 (0.78); FEV1/FVC: −0.68 (1.20); FEF25-75: −0.99 (1.02)] were similar to children with ‘wheeze.’

## Discussion

Our population-based study found children reporting dry night cough without wheeze (‘cough’) are distinct from children with ‘wheeze’ or ‘wheeze and cough’ in terms of clinical characteristics, family history, FeNO, lung function, healthcare use and asthma diagnosis and treatment. Only a small subgroup (7%) of children reporting ‘cough’ were diagnosed with asthma and treated with inhalers; their features were similar to children with ‘wheeze.’

### Comparison with other studies

Other population-based studies found children who cough without wheeze differed from children who wheeze [14, 20–23, 29]. A study of 8–13 year olds in Scotland found a personal history of eczema and parental history of asthma more common among children with night cough than asymptomatic children, but less common among children with wheeze-related symptoms [20]. We did not find the same, possibly because they asked about cough of extended duration (≥4 weeks). In Finland, prevalence of parental asthma was higher among children with dry night cough alone than asymptomatic children but lower than among children with wheeze [23]. As we, this study, reported that respiratory symptoms triggered by pollen, pets, and house dust were less common in children with cough compared to children with wheeze.

FeNO has not been described in studies comparing children with cough and wheeze. Although spirometry was measured in some population-based studies, it was mostly among preschool children [22, 24, 25]. Only one study included schoolchildren and found the maximal mid-expiratory flow higher among children with dry night cough when compared with children with wheeze, while there were no differences in FVC and FEV1 [23]. Further studies are needed to determine whether our findings of higher FVC among children with ‘cough’ than ‘asymptomatic’ children was chance or a true phenomenon, perhaps via a training effect of vital capacity and full expiration among children with recurrent cough.

Since hardly any children with ‘cough’ in our study visited the hospital emergency unit and no children were hospitalised, it means their symptoms–while bothersome–were not alarming. Similarly, in a study of children aged 6–12 years in Australia, no hospital admissions were reported among children with cough only, although they used a more stringent definition of cough (lasting more than 3 weeks without a cold or flu)[29].

### Interpretation and implication of findings

The new ERS clinical guidelines for diagnosing asthma among schoolchildren recommend wheeze as requisite symptom for asthma (*i.e.* not diagnosing asthma for children only reporting cough) and measuring FeNO and lung function in the asthma diagnostic work-up [39], which our results support. Indeed, we found most children who report dry night cough without wheeze (‘cough’) do not have typical features of asthma. We found no evidence of bronchial obstruction or eosinophilic airway inflammation and no association with family history of asthma or a personal history of hay fever or eczema. However, local healthcare providers diagnosed asthma and prescribed asthma inhalers for a small proportion (7%) of children with dry night cough alone. We offer two explanations: asthma characterised by cough without wheeze may exist, yet it is rare; or children’s wheeze was unrecognized and unreported by parents. In the UK, many parents incorrectly understand the word wheeze [40, 41], which might be similar among a German-speaking population such as ours since the German language lacks a specific word for wheeze.

We found ‘cough’ was associated with lower socioeconomic status, more frequent colds and ENT problems, such as rhinitis and snoring. Lower socioeconomic status is known to be associated with exposure to infectious agents and decreased host resistance to infection [42]. The association of cough with the report of ENT-related complaints, such as rhinitis and snoring, may be explained by several mechanisms, such as stimulating pharyngeal cough receptors by post-nasal drip, hyper-responsive cough receptors, or mouth breathing due to nasal obstructions, leading to reduced filtration, humidification, and warming of inhaled air [43]. In a study of 103 children with upper airway cough syndrome suffering from chronic cough, adenoidal hypertrophy was the main cause for pre-school children whereas allergic or non-allergic rhinitis caused chronic cough for the majority of school-aged children [44]. Our findings suggest investigating children with isolated cough for differential diagnoses, particularly upper respiratory problems.

As a novel aspect, our study compared children reporting ‘wheeze and cough’ with children reporting only ‘wheeze.’ We found higher FeNO and more parental asthma among children with ‘wheeze,’ suggesting they might have an asthma phenotype linked closely to atopy. Children with ‘wheeze and cough’ frequently reported upper respiratory symptoms (otitis media, rhinitis) and non-allergic triggers, such as cold air and temperature changes, suggesting some of these children may have two independent problems (*i.e.* cough unrelated to coexisting asthma). When two diseases are common, by chance a proportion of patients have both.

### Strengths and limitations

Our study has several strengths. It is large, population-based and representative of children in the community with dry night cough or wheeze. We collected details of accompanying symptoms and medical histories and measured FeNO and spirometry—two objective traits related to asthma. We also distinguished four mutually exclusive groups, which allowed for investigating differences between children with both night cough and wheeze compared with those with wheeze alone. We gathered data about healthcare utilisation and treatment by local physicians. Limitations of our study include lack of information about night cough duration, objective recordings of respiratory sounds, and tests assessing allergy, bronchial responsiveness or cough receptor sensitivity, which are not possible to collect for school-based studies.

## Conclusion

Our findings align with the new ERS guidelines for diagnosing asthma among schoolchildren. The new ERS guidelines state children with only cough as a symptom are unlikely to have asthma and differential diagnoses, particularly upper respiratory problems, should be investigated. However, a minority of children (7%) shares characteristics with children who wheeze. These may have a variant form of asthma, and they could benefit from an asthma diagnostic work-up and treatment.

## Supporting information

STROBE Checklist

## Data Availability

The anonymised dataset used for this study is available from the corresponding author upon reasonable request and if the study group members consent.

## Acknowledgements

We thank the LUIS study group, the school staff, the LUIS study fieldworkers, the parents and children who participated in our study and Kristin Marie Bivens, PhD for her editorial contribution.

## LUIS study group members

Alexander Moeller, Jakob Usemann (Department of Respiratory Medicine, University Children’s Hospital Zurich and Childhood Research Centre, University of Zurich, Switzerland); Philipp Latzin, Florian Singer and Johanna Kurz (Paediatric Respiratory Medicine, Children’s University Hospital of Bern, University of Bern, Switzerland); Claudia E. Kuehni, Rebeca Mozun, Cristina Ardura-Garcia, Myrofora Goutaki, Eva S.L. Pedersen and Maria Christina Mallet (Institute of Social and Preventive Medicine, University of Bern, Switzerland); Kees de Hoogh (Swiss Tropical and Public Health Institute, Basel, Switzerland).

## Funding

Lunge Zürich, Switzerland funded data collection and the Swiss National Science Foundation (SNF Grant: 320030_182628) funded our analysis.

## Conflicts of interest

Mallet MC, Mozun R, Ardura-Garcia C, Pedersen ESL, Jurca M, Kuehni CE have nothing to disclose. Latzin P reports personal fees from OM pharma, Polyphor, Santhera, Vertex, Vifor, Sanofi Aventis and grants from Vertex—all outside the submitted work. Moeller A reports personal fees from Vertex outside the submitted work.

---

**Figure S1:**
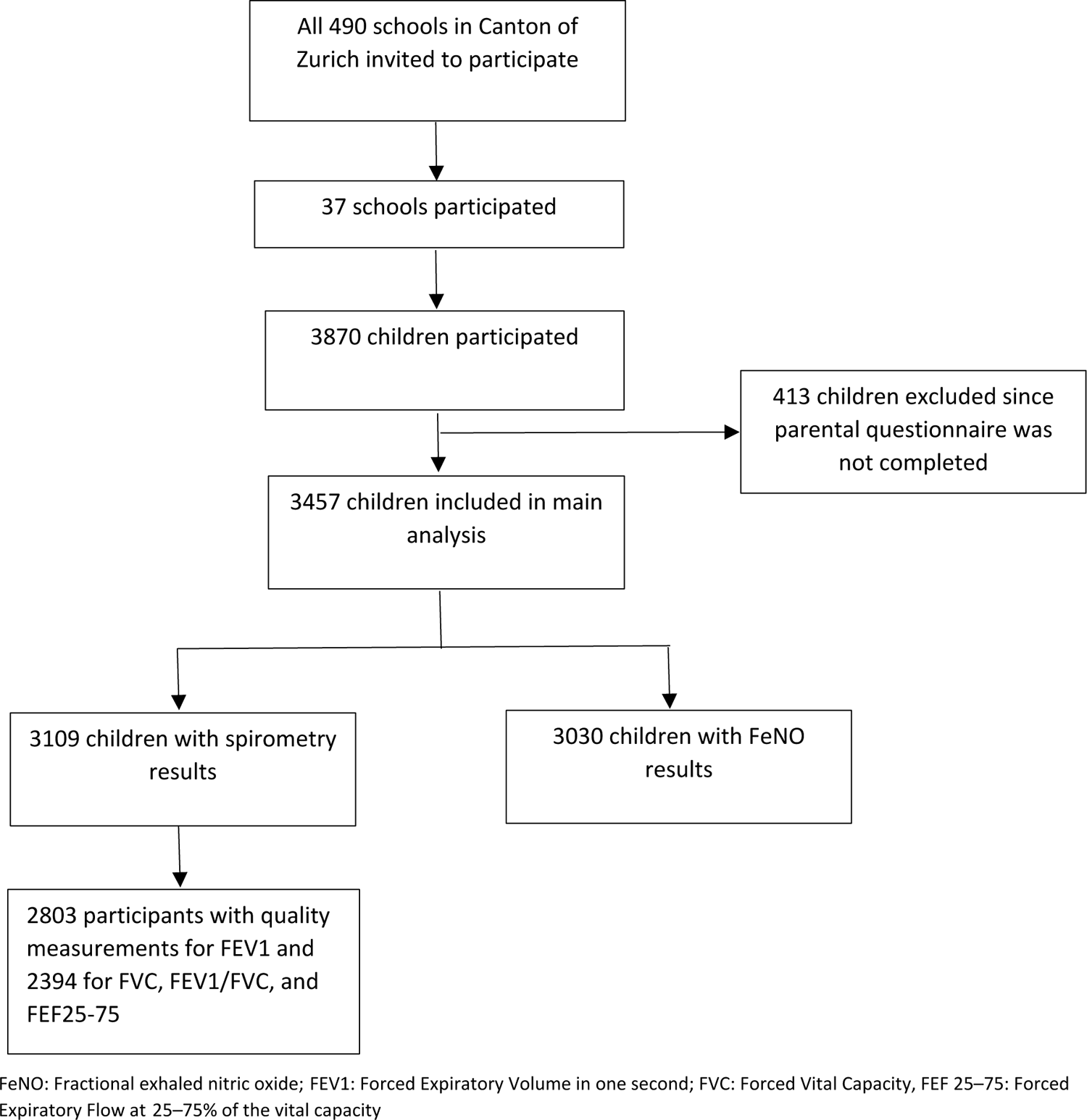
Flowchart of included participants

**Figure S2A:**
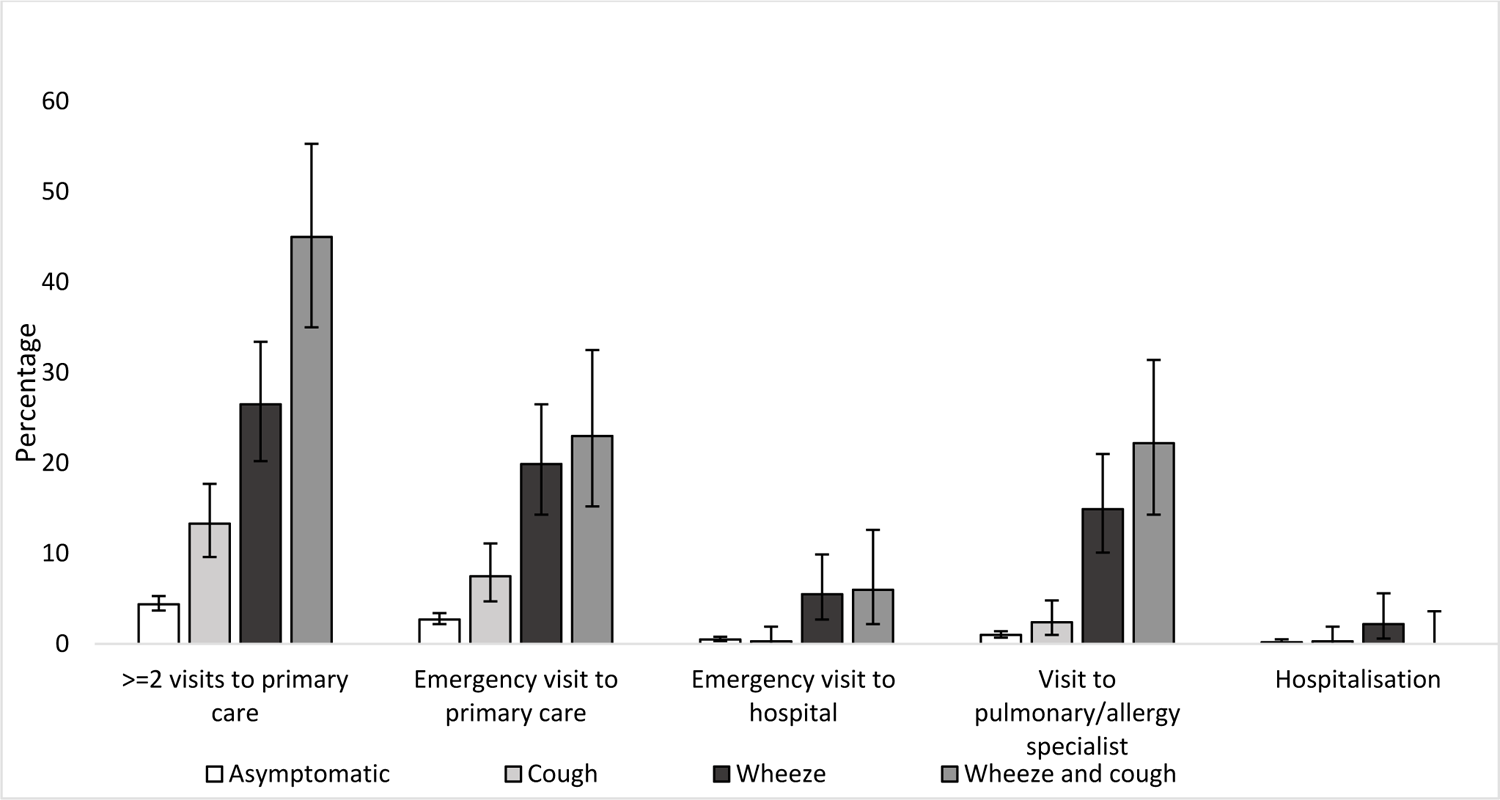
Healthcare utilisation for respiratory complaints in the past 12 months among asymptomatic children (n=2882), children with cough (n=294), wheeze (n=181), and wheeze and cough (n=100)

**Figure S2B:**
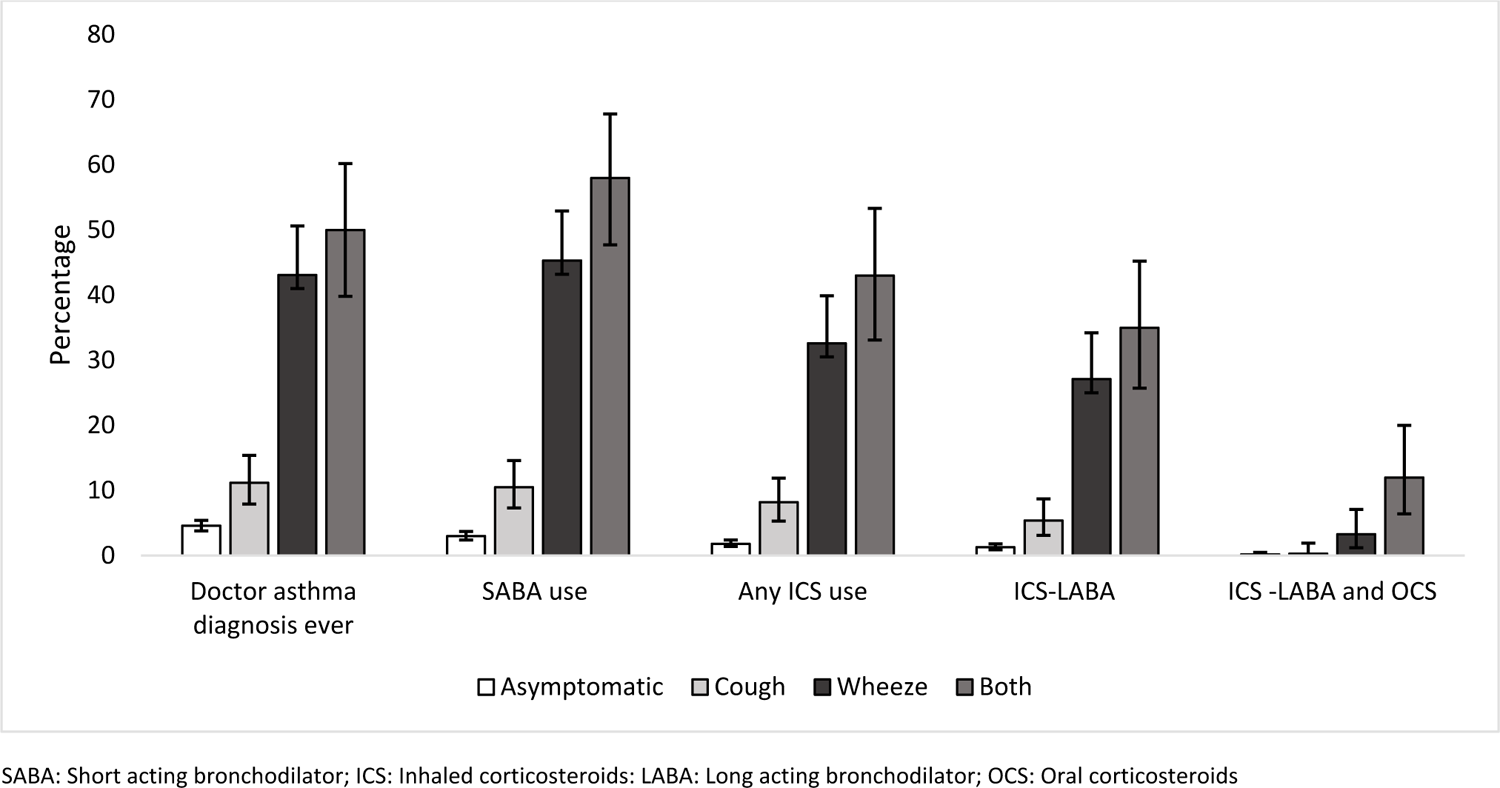
Frequency of physician diagnosis of asthma ever and asthma treatment in the past 12 months among asymptomatic children (N=2882), children with cough (N=294), wheeze (N=181), and wheeze and cough (N=100)

**Table S1:** Wording of questions, question source, answer categories, and missing values

| Variable | Question | Question source | Answer categories, N (%) | Comments |
| --- | --- | --- | --- | --- |
| **OUTCOME** |  |  |  |  |
| Dry night cough | In the last 12 months, has your child had a dry cough at night apart from a cough associated with a cold or a chest infection? | ISAAC | No: 3017 (87)<br>Yes: 394 (11)<br>Missing: 46 (1) | Missing variable recoded as no |
| Wheeze | Did your child have wheezing or whistling in the chest in the past 12 months? | ISAAC | No: 3118 (90)<br>Yes: 281 (8)<br>Missing: 58 (2) | Missing variable recoded as no |
| **EXPOSURES** |  |  |  |  |
| Sex |  |  | Male: 1736 (50)<br>Female: 1719 (50)<br>Missing: 2 (0) |  |
| Place of birth | In which country was the child born? | LRC | Switzerland: 3042 (88)<br>Other: 391 (11)<br>Missing: 24 (1) |  |
| Swiss socioeconomic position (SEP) index | Based on neighbourhood/addresses. The geocode of participant addresses were matched with the nearest geocode in the Swiss-SEP dataset, which was developed by the Swiss National Cohort (SNC). |  | Continuous value from 0 (lowest) to 100 (highest) | If participant addresses were missing, we assigned the median Swiss-SEP among participants from the same school |
| Urbanisation degree | Defined by the Swiss Federal Office of Statistics classification |  | Large urban : 1674 (48)<br>Small urban: 1538 (44)<br>Rural areas: 245 (7)<br>Missing: 0 (0) | Small urban and rural areas combined into one category |
| Household pets | Do you keep any household pets? | LRC | Yes: 1955 (57)<br>No: 1469 (42)<br>Missing: 33 (1) |  |
| Number of siblings | How many children live in your household? | LRC | Continuous variable organised into 3 categories:<br>0 to 1: 569 (16)<br>2 to 3: 2537 (73)<br>>3: 259 (7)<br>Missing: 92 (3) |  |
| Presence of older siblings | How many children are older than the questioned child? |  | No: 1647 (48)<br>Yes: 1552 (45)<br>Missing: 258 (7) | Variable dichotomised as no (if no children older) and yes (if at least one children older) |
| Maternal smoking | Does the mother smoke? |  | No: 2856 (83)<br>Yes (outdoor and also in apartment): 567 (17)<br>Missings:30 (1) | All yes categories combined |
| Paternal smoking | Does the mother smoke? |  | No: 2644 (76)<br>Yes (outdoor and also in apartment): 757 (22)<br>Missing: 56 (2) | All yes categories combined |
| Body Mass Index (BMI) z-scores | Height and weight measured in the bus without wearing shoes |  | Continuous; BMI z-scores calculated using World Health Organisation reference tables<br>Missing: 96 (3) |  |
| Frequency of colds | How many colds did your child have in the last 12 months? | LRC | None: 224 (6)<br>1 to 3: 2566 (74)<br>4 to 6: 499 (14)<br>7 to 9: 67 (2)<br>10 or more: 21 (1)<br>Missing: 80 (2) | Missing variable recoded as 0. We created a binary category: 0–3 and >3 |
| Family history of asthma | Does the child's mother or father suffer from asthma or does she/he need an asthma spray to inhale? |  | No, neither of them: 2923 (85)<br>Yes, the mother: 268 (8)<br>Yes, the father: 196 (6)<br>Yes, both: 22 (1)<br>Missing: 48 (1) | Variable dichotomised as no and yes (mother, father, or both) |
| Family history of chronic cough | Does the child's mother or father suffer from chronic cough or bronchitis? |  | No, neither of them: 3085 (89)<br>Yes, the mother: 80 (2)<br>Yes, the father: 94 (3)<br>Yes, both: 6 (0)<br>Missing: 192 (6) | Variable dichotomised as no and yes (mother, father, or both) |
| Personal history of hay fever | Has your child ever had hay fever? | ISAAC | No: 2612 (76)<br>Yes: 766 (22)<br>Missing: 79 (2) | Missing variable recoded as no |
| Personal history of eczema | Has a doctor told you that your itchy rash in the past 12 months was eczema? |  | No: 3131 (91)<br>Yes: 249 (7)<br>Missing: 77 (2) | Missing variable recoded as no |
| Rhinitis | In the past 12 months, has your child had a problem with sneezing, or a runny, or a blocked nose when he/she did not have a cold or flu? | ISAAC | No: 2369 (69)<br>Yes: 1048 (30)<br>Missing: 40 (1) | Missing variable recoded as no |
| Otitis media | Did your child have middle ear infection in the last 12 months? | LRC | No: 3082 (89)<br>Yes, once: 268 (8)<br>Yes, more than once: 65 (2)<br>Missing: 42 (1) | Missing variable recoded as no<br>Variable dichotomised as no and yes |
| Snoring | How often did your child snore in the last 12 months | LRC | No snoring: 1938 (56)<br>Only during colds: 527 (15)<br>Also without colds: 725 (21)<br>Almost every night: 163 (5)<br>Missing: 104 (3) | Missing variable recorded as no<br>Snoring was defined as those who reported snoring also without colds and almost every night |
| History of adeno-tonsillectomy | Did your child have tonsillectomy? |  | No: 3233 (94)<br>Yes: 188 (5)<br>Missing: 36 (1) | Missing variable recoded as no |

ISAAC: international study of asthma and allergies in childhood, LRC: Leicester Respiratory Cohort

**Table S2:**
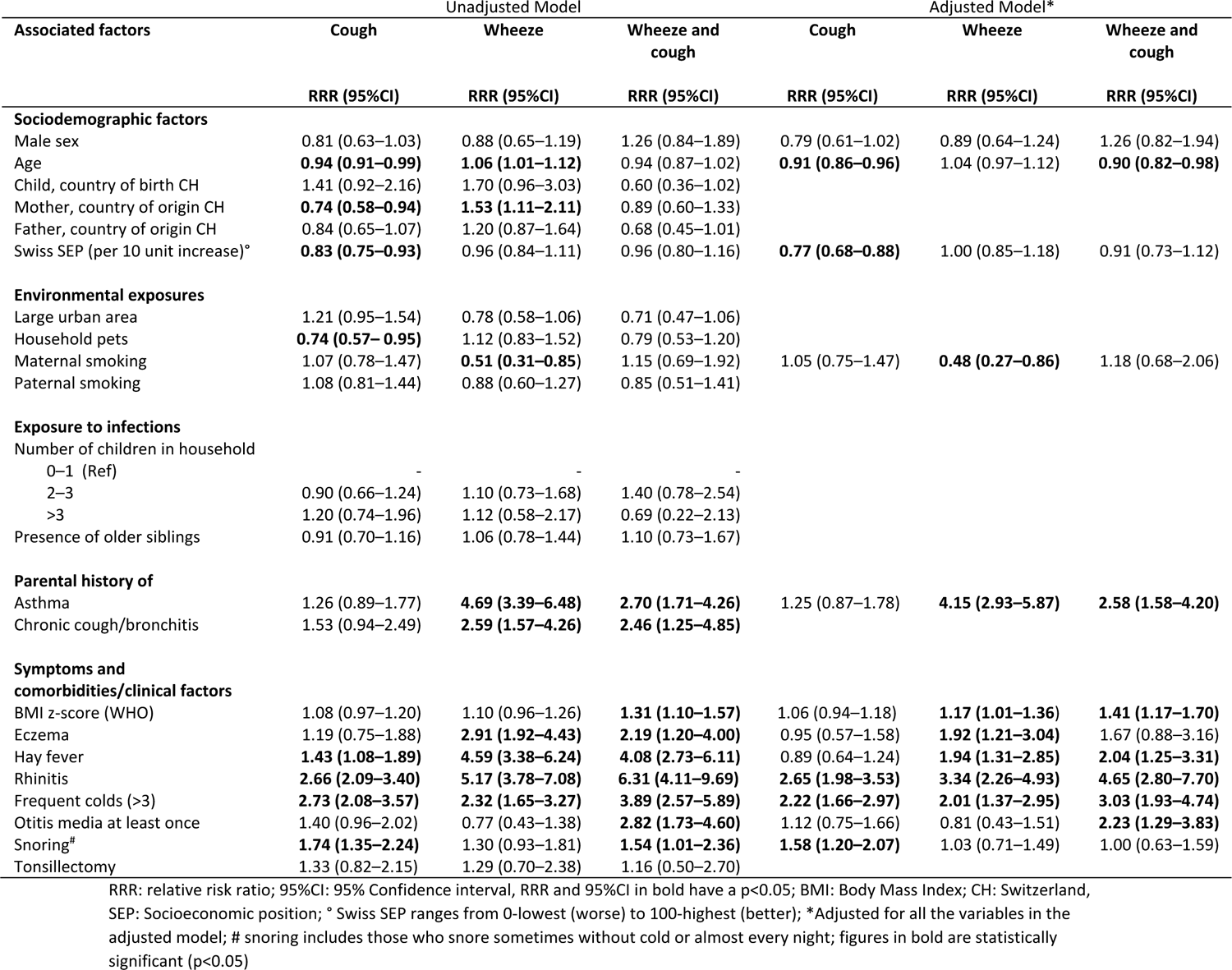
Factors associated with cough (N=294), wheeze (N=181), and wheeze and cough (N=100) compared with asymptomatic (N=2882) (reference group)

**Table S3:**
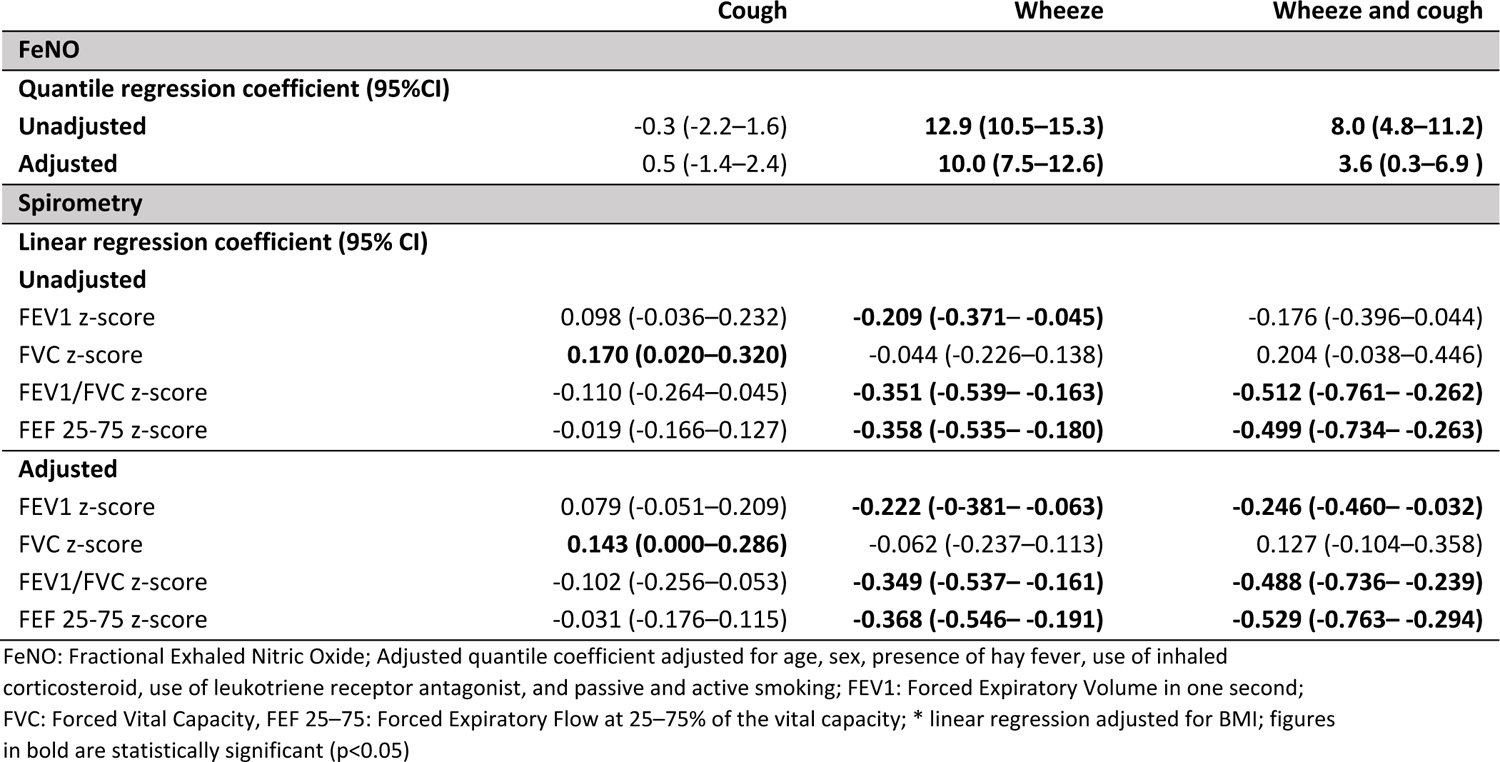
Association of FeNO and spirometry z-scores for children with cough, wheeze, and wheeze and cough compared with asymptomatic children (reference group)

|  | Cough | Wheeze | Wheeze and cough |
| --- | --- | --- | --- |
| **FeNO** |  |  |  |
| **Quantile regression coefficient (95%CI)** |  |  |  |
| **Unadjusted** | -0.3 (-2.2–1.6) | **12.9 (10.5–15.3)** | **8.0 (4.8–11.2)** |
| **Adjusted** | 0.5 (-1.4–2.4) | **10.0 (7.5–12.6)** | **3.6 (0.3–6.9)** |
| **Spirometry** |  |  |  |
| **Linear regression coefficient (95% CI)** |  |  |  |
| **Unadjusted** |  |  |  |
| FEV1 z-score | 0.098 (-0.036–0.232) | **-0.209 (-0.371– -0.045)** | -0.176 (-0.396–0.044) |
| FVC z-score | **0.170 (0.020–0.320)** | -0.044 (-0.226–0.138) | 0.204 (-0.038–0.446) |
| FEV1/FVC z-score | -0.110 (-0.264–0.045) | **-0.351 (-0.539– -0.163)** | **-0.512 (-0.761– -0.262)** |
| FEF 25-75 z-score | -0.019 (-0.166–0.127) | **-0.358 (-0.535– -0.180)** | **-0.499 (-0.734– -0.263)** |
| **Adjusted** |  |  |  |
| FEV1 z-score | 0.079 (-0.051–0.209) | **-0.222 (-0.381– -0.063)** | **-0.246 (-0.460– -0.032)** |
| FVC z-score | **0.143 (0.000–0.286)** | -0.062 (-0.237–0.113) | 0.127 (-0.104–0.358) |
| FEV1/FVC z-score | -0.102 (-0.256–0.053) | **-0.349 (-0.537– -0.161)** | **-0.488 (-0.736– -0.239)** |
| FEF 25-75 z-score | -0.031 (-0.176–0.115) | **-0.368 (-0.546– -0.191)** | **-0.529 (-0.763– -0.294)** |

FeNO: Fractional Exhaled Nitric Oxide; Adjusted quantile coefficient adjusted for age, sex, presence of hay fever, use of inhaled corticosteroid, use of leukotriene receptor antagonist, and passive and active smoking; FEV1: Forced Expiratory Volume in one second; FVC: Forced Vital Capacity, FEF 25–75: Forced Expiratory Flow at 25–75% of the vital capacity; \* linear regression adjusted for BMI; figures in bold are statistically significant (p<0.05)

**Table S4:**
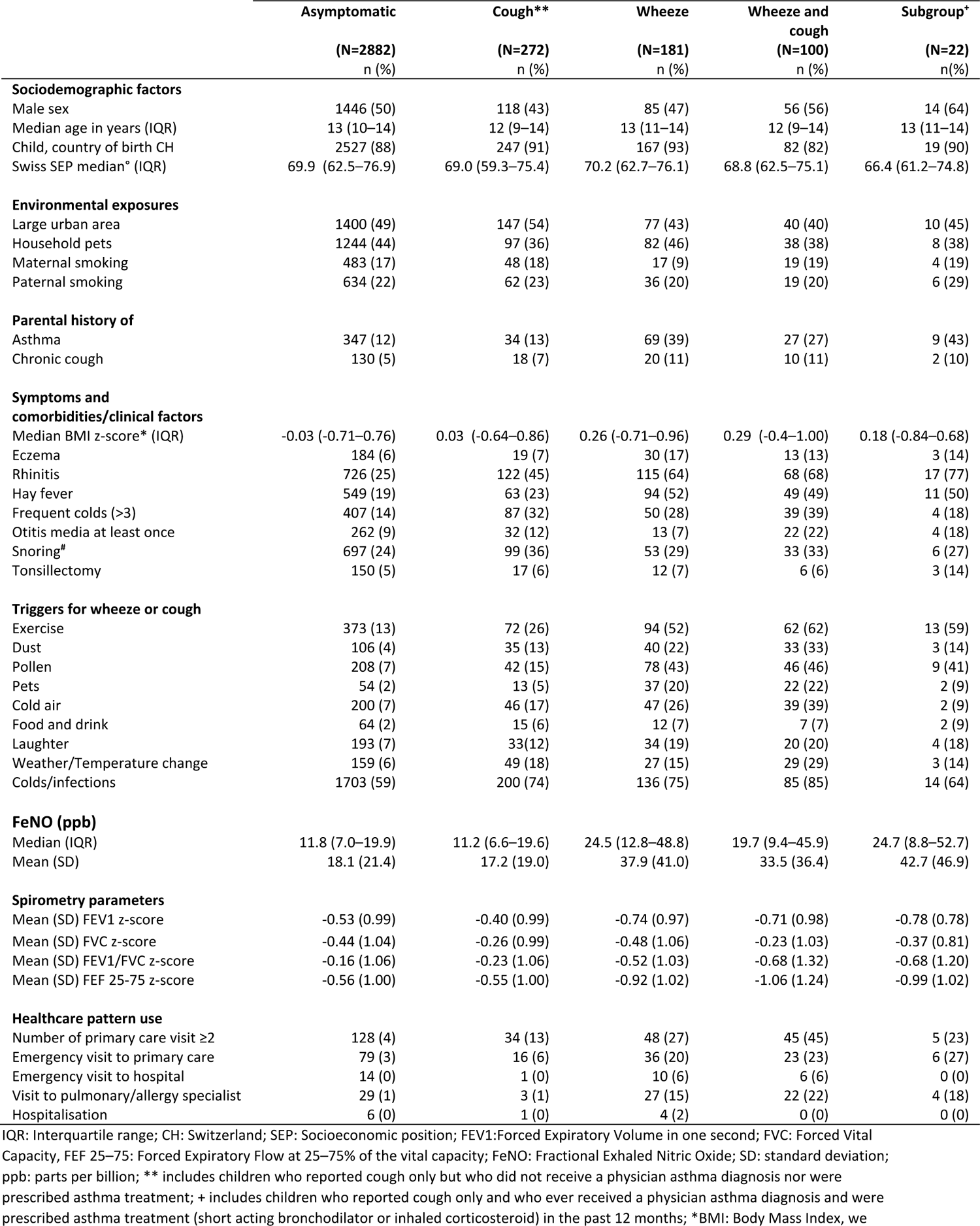
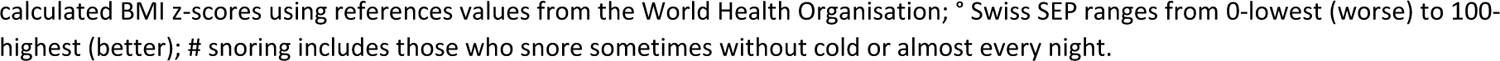
Reported characteristics and measured traits of a subgroup of children with cough who were ever diagnosed as asthma and received asthma treatment in the past 12 months (n=22) compared with asymptomatic children, the remaining children with cough, and children with wheeze and wheeze and cough.

|  | Asymptomatic | Cough** | Wheeze | Wheeze and cough | Subgroup<sup>+</sup> |
| --- | --- | --- | --- | --- | --- |
|  | (N=2882)<br>n (%) | (N=272)<br>n (%) | (N=181)<br>n (%) | (N=100)<br>n (%) | (N=22)<br>n(%) |
| **Sociodemographic factors** |  |  |  |  |  |
| Male sex | 1446 (50) | 118 (43) | 85 (47) | 56 (56) | 14 (64) |
| Median age in years (IQR) | 13 (10–14) | 12 (9–14) | 13 (11–14) | 12 (9–14) | 13 (11–14) |
| Child, country of birth CH | 2527 (88) | 247 (91) | 167 (93) | 82 (82) | 19 (90) |
| Swiss SEP median<sup>o</sup> (IQR) | 69.9 (62.5–76.9) | 69.0 (59.3–75.4) | 70.2 (62.7–76.1) | 68.8 (62.5–75.1) | 66.4 (61.2–74.8) |
| **Environmental exposures** |  |  |  |  |  |
| Large urban area | 1400 (49) | 147 (54) | 77 (43) | 40 (40) | 10 (45) |
| Household pets | 1244 (44) | 97 (36) | 82 (46) | 38 (38) | 8 (38) |
| Maternal smoking | 483 (17) | 48 (18) | 17 (9) | 19 (19) | 4 (19) |
| Paternal smoking | 634 (22) | 62 (23) | 36 (20) | 19 (20) | 6 (29) |
| **Parental history of** |  |  |  |  |  |
| Asthma | 347 (12) | 34 (13) | 69 (39) | 27 (27) | 9 (43) |
| Chronic cough | 130 (5) | 18 (7) | 20 (11) | 10 (11) | 2 (10) |
| **Symptoms and comorbidities/clinical factors** |  |  |  |  |  |
| Median BMI z-score* (IQR) | -0.03 (-0.71–0.76) | 0.03 (-0.64–0.86) | 0.26 (-0.71–0.96) | 0.29 (-0.4–1.00) | 0.18 (-0.84–0.68) |
| Eczema | 184 (6) | 19 (7) | 30 (17) | 13 (13) | 3 (14) |
| Rhinitis | 726 (25) | 122 (45) | 115 (64) | 68 (68) | 17 (77) |
| Hay fever | 549 (19) | 63 (23) | 94 (52) | 49 (49) | 11 (50) |
| Frequent colds (>3) | 407 (14) | 87 (32) | 50 (28) | 39 (39) | 4 (18) |
| Otitis media at least once | 262 (9) | 32 (12) | 13 (7) | 22 (22) | 4 (18) |
| Snoring<sup>#</sup> | 697 (24) | 99 (36) | 53 (29) | 33 (33) | 6 (27) |
| Tonsillectomy | 150 (5) | 17 (6) | 12 (7) | 6 (6) | 3 (14) |
| **Triggers for wheeze or cough** |  |  |  |  |  |
| Exercise | 373 (13) | 72 (26) | 94 (52) | 62 (62) | 13 (59) |
| Dust | 106 (4) | 35 (13) | 40 (22) | 33 (33) | 3 (14) |
| Pollen | 208 (7) | 42 (15) | 78 (43) | 46 (46) | 9 (41) |
| Pets | 54 (2) | 13 (5) | 37 (20) | 22 (22) | 2 (9) |
| Cold air | 200 (7) | 46 (17) | 47 (26) | 39 (39) | 2 (9) |
| Food and drink | 64 (2) | 15 (6) | 12 (7) | 7 (7) | 2 (9) |
| Laughter | 193 (7) | 33(12) | 34 (19) | 20 (20) | 4 (18) |
| Weather/Temperature change | 159 (6) | 49 (18) | 27 (15) | 29 (29) | 3 (14) |
| Colds/infections | 1703 (59) | 200 (74) | 136 (75) | 85 (85) | 14 (64) |
| **FeNO (ppb)** |  |  |  |  |  |
| Median (IQR) | 11.8 (7.0–19.9) | 11.2 (6.6–19.6) | 24.5 (12.8–48.8) | 19.7 (9.4–45.9) | 24.7 (8.8–52.7) |
| Mean (SD) | 18.1 (21.4) | 17.2 (19.0) | 37.9 (41.0) | 33.5 (36.4) | 42.7 (46.9) |
| **Spirometry parameters** |  |  |  |  |  |
| Mean (SD) FEV1 z-score | -0.53 (0.99) | -0.40 (0.99) | -0.74 (0.97) | -0.71 (0.98) | -0.78 (0.78) |
| Mean (SD) FVC z-score | -0.44 (1.04) | -0.26 (0.99) | -0.48 (1.06) | -0.23 (1.03) | -0.37 (0.81) |
| Mean (SD) FEV1/FVC z-score | -0.16 (1.06) | -0.23 (1.06) | -0.52 (1.03) | -0.68 (1.32) | -0.68 (1.20) |
| Mean (SD) FEF 25-75 z-score | -0.56 (1.00) | -0.55 (1.00) | -0.92 (1.02) | -1.06 (1.24) | -0.99 (1.02) |
| **Healthcare pattern use** |  |  |  |  |  |
| Number of primary care visit ≥2 | 128 (4) | 34 (13) | 48 (27) | 45 (45) | 5 (23) |
| Emergency visit to primary care | 79 (3) | 16 (6) | 36 (20) | 23 (23) | 6 (27) |
| Emergency visit to hospital | 14 (0) | 1 (0) | 10 (6) | 6 (6) | 0 (0) |
| Visit to pulmonary/allergy specialist | 29 (1) | 3 (1) | 27 (15) | 22 (22) | 4 (18) |
| Hospitalisation | 6 (0) | 1 (0) | 4 (2) | 0 (0) | 0 (0) |

IQR: Interquartile range; CH: Switzerland; SEP: Socioeconomic position; FEV1: Forced Expiratory Volume in one second; FVC: Forced Vital Capacity, FEF 25–75: Forced Expiratory Flow at 25–75% of the vital capacity; FeNO: Fractional Exhaled Nitric Oxide; SD: standard deviation; ppb: parts per billion; \*\* includes children who reported cough only but who did not receive a physician asthma diagnosis nor were prescribed asthma treatment; + includes children who reported cough only and who ever received a physician asthma diagnosis and were prescribed asthma treatment (short acting bronchodilator or inhaled corticosteroid) in the past 12 months; \*BMI: Body Mass Index, we
calculated BMI z-scores using references values from the World Health Organisation; ° Swiss SEP ranges from 0-lowest (worse) to 100-highest (better); # snoring includes those who snore sometimes without cold or almost every night.

## Notes

### Author Declarations

The ethics committee of the canton of Zurich approved the study (KEK-ZH-Nr (Kantonalen Ethikkommission Zürich): 2014-0491)

